# Implementation and Impact of a Diversity Supplement Repository

**DOI:** 10.1101/2025.01.24.25321089

**Authors:** Maryam Gholami, Eva Kintzer, Mitchell Wong, Davey Smith, Colin Depp

## Abstract

The National Institutes of Health (NIH) diversity supplements represent an opportunity to enhance diversity in the biomedical research workforce. Despite their potential impact, practical barriers prevent effective use of these resources. The Altman Clinical and Translational Research Institute (ACTRI) at the University of California San Diego (UCSD) developed and implemented of an institutional repository and support system to improve diversity supplement applications. The centralized repository and support system incorporated three strategies: (1) a secure web-based repository housing successful examples of supplements, (2) match making for diversity supplements and mentors, and (3) web-based resources for potential applicants. The repository was implemented in June 2021 and includes 51 supplement samples across 14 divisions of NIH as of November 2024. The repository has been accessed by 51 potential applicants since implementation in 2021. Few investigators have requested match making. Early indicators show a doubling in diversity supplement applications compared to pre-implementation period at UCSD. We outlined our approach to addressing some of the barriers in diversity supplement applications, which could provide a model for other institutions. Pending solutions to some of the lessons learned, coordinated efforts aimed at diversity supplements could be a practical approach toward a more diverse biomedical research workforce.

## INTRODUCTION

The National Institutes of Health (NIH) Diversity Supplement program represents a significant investment in enhancing diversity in the biomedical research workforce. This administrative supplement mechanism provides additional funding to eligible NIH research grants to support individuals from underrepresented groups in biomedical research (1). The program is implemented across 26 different NIH institutes. These supplements serve multiple career stages, from graduate students to early-career faculty, providing crucial support for research training and career development in biomedical sciences. Hill et al. (2) documented the distribution of NIH Diversity Supplement Awards across various administering institutes, highlighting both the reach and impact of the program, and that the program is not fully utilized. Recent reports have identified this program as an underutilized opportunity to improve diversity in health sciences research (3). In this paper, we detail experiences and lessons learned in learned in UCSD’s approach to increasing diversity supplements.

Studies have documented substantial research productivity outcomes among diversity supplement recipients (1). The research output includes increased publication rates following their supplement period, with citation rates demonstrating the quality and impact of their work. Moreover, program evaluation has evidenced enhanced research productivity among supplement recipients in areas addressing health disparities and underserved populations (1). This finding aligns with the program’s goal of fostering diverse perspectives in biomedical research. Yorke et al. (4) specifically examined experiences in radiation oncology, finding that the diversity supplement program effectively supports trainees’ transition to independent positions while enhancing their research skills and scientific competencies. Gandhi & Johnson (5) emphasized the importance of effective mentoring in achieving these educational outcomes, suggesting that mentor training programs can further enhance the success of diversity supplements. This mentorship component is particularly critical as it shapes the experiences of both mentees and mentors throughout the supplement period.

The biomedical research funding landscape shows persistent racial and ethnic disparities (6). While the root causes are complex, these disparities emerge early in researchers’ careers, affecting their advancement to higher academic positions, tenure, and funding opportunities (4). This pattern underscores the critical need for early-career interventions to promote equity in research funding. An analysis of NIH diversity supplement awards from 2005 to 2020 revealed significant underuse of this funding mechanism across medical schools, regardless of their NIH funding status (7). Despite the substantial growth in awards from 17 to 270 per year between 2012 and 2020 (41.3% annual growth rate), only 2.12% of R01 grants have an associated diversity supplement, indicating a missed opportunity for developing diverse faculty. The considerable variation in use among medical schools (ranging from 4 to 56 supplements per school) and the percentage of R01 grants with supplements (ranging from 0.79% to 4.5%) indicates heterogeneity in institutional approaches to pursuing these opportunities. This variation, combined with the fact that three NIH Top 40 medical schools received no R01-associated diversity supplements in 2020, highlights the need for enhanced institutional support and infrastructure to streamline the application process. The data suggests that medical schools are missing crucial opportunities to mentor, retain, and develop faculty from underrepresented groups, particularly given that 70.8% (n = 983) of all R01-associated diversity supplements went to Top 40 institutions, indicating a concentration of resources that could be more broadly distributed to enhance diversity across the biomedical research workforce.

Research by David et al. (1) revealed several barriers that contribute to the underuse of diversity supplements. Some barriers include identified low program awareness on the part of both potential applicants and principal investigators remaining unaware of these opportunities. The application process itself presents significant complexity and variability across NIH institutes, with requirements that can be daunting to navigate for both mentees and mentors. Additionally, the form level of administrative support varies considerably across NIH institutions. The CTSA network provides a unique platform to create, collaborate, and disseminate resources to address observed gaps in diversity supplements.

Recognizing an unmet need at University of California San Diego (UCSD), the Altman Clinical and Translational Research Institute (ACTRI) in 2021 set out to create a set of resources for local and regional investigators and potential awardees to facilitate diversity supplements. We detail these processes and resources here, including collaboration with other CTSA hubs, in hopes that this information and the lessons learned could be useful for other institutions.

## METHODS

### Overview

The initial aims of the diversity supplement program were to 1) create a repository of successful examples, 2) develop a match making function wherein potential applicants could be linked with potential mentors and parent grants, 3) generate guides to detail the process of applying for a supplement, and 4) foster an opt-in network for potential PIs and awardees to receive notification of specific funding opportunities.

### Institutional Collaborations in Repository Development

To establish a secure repository for successful grant applications, we partnered with two CTSA hubs. UC Irvine introduced us to FlowPaper, a cloud-based solution preventing document copying, while UCLA, which pioneered grant repositories among western CTSA hubs, shared their expertise in document security and distribution through password-protected links. This collaboration led to a shared approach for growing both institutions’ diversity supplement repositories while maintaining document security and investigator privacy.

### Identifying and Recruitment Process

#### Survey of the rate of diversity supplements and identified awardees at UCSD

To assess UCSD’s diversity supplement landscape, we developed a systematic approach for identifying awarded supplements using NIH Reporter and eRA Commons databases. Initial identification through eRA Commons (2018-2021) revealed 48 diversity awards among 263 supplement awards reviewed. We refined our methodology using NIH Reporter’s advanced search function with specific parameters including UCSD organizational variations, fiscal years, funding agencies, and diversity-specific FOA numbers (PA-05-015 through PA-21-071).

Analysis of 2019 data showed 12 funded diversity supplements against over 700 potentially eligible grants (R or U awards with remaining funding). To track and manage outreach, we created a database using Monday.com that incorporated validated award information and contact status. This improved identification process allowed for more accurate tracking of diversity supplement utilization at UCSD.

#### Outreach to PIs

We developed an email template to send to identified investigator awardees, where we highlighted UCSD’s diversity supplement underutilization (only 12 awards in 2019) despite high success rates. The email template focused on three main objectives: creating a repository of successful applications to simplify the process, establishing UCSD as a leader in diversity supplement initiatives, and gathering best practices from successful recipients. The aim was to both collect successful applications and develop a supportive network for future applicants.

#### Recruitment and consenting approach for PI and awardee recipient

Once initial consent was received from the awardee lead or contact investigator, we asked for contact information to request student trainee consent. When professional relationships remained intact, investigators often facilitated trainee consent. Document sharing preferences were established during consent, with options ranging from UCSD-only access to broader institutional sharing. From there, follow-up communication with the investigator’s administrative assistant or trainee was usually required to receive an actual sample document. Applications were excluded when trainees were unresponsive or when investigator-trainee relationships had deteriorated post-award. We share approved successful samples mutually with each other and currently there are no other institutional collaborations besides UCLA.

### Data Collection and Storage

#### Document preparation and safeguarding

We prepared received sample documents by applying redactions and other safeguarding measures. Using Adobe Acrobat, we redacted all trainee personal information, unless instructed otherwise, including but not limited to names and initials, login username and typo names as well as signature, date of birth, SSN, and phone number if applicable. Some contributors preferred to self-redact private information before submitting and a small number preferred no redactions sending in their document as is. Newer awardees often preferred full redaction of their documents removing certain sections of the application relating to sensitive science, novel discoveries, and pending patents. They also tended to deny the broader sharing option to which we approached investigators about their openness to our team circling back in a year or two to ask again. Our quality control process included secondary review of redactions, followed by watermarking all pages with distribution restrictions. Title pages were applied to the front of finalized documents. As a final security measure, we relied on FlowPaper, a cloud-based hosting solution that converts documents into non-downloadable, copy-protected files with password protection. This platform allows for unlimited publications with up to 50 shareable web-based links through the Enterprise Zine subscription. Changing subscription tier is possible anytime without impacting file versions shared on the prior tier level. These publication links are password protected during the process of Cloud-hosted conversion using FlowPaper.

#### Accessibility of resources

UCSD uses a secured approved share drive system allowing storing and access to documents, files and other shared resources securely. Our diversity supplement repository is a collection of document file links that is set up as a table of contents including PI name, project title, department, IC or sponsor agency, campus, year of award and level of trainee. We have two separate running shareable repositories: 1) one only approved for sharing with the UCSD community and its affiliates and 2) one approved for sharing outside of UCSD with anyone whose email address is institution affiliated. Repository access requires a two-step verification process: completion of a Monday.com request form and submission of a legally binding attestation agreement signed through DocuSign Power Form confirming acceptance of usage terms.

### Dissemination Strategies

#### Webinar

UCSD partnered with San Diego State University to host a Diversity Supplements webinar in May 2022, attracting over 100 registrants. The event featured a panel of leaders from five NIH institutes (NIMH, NCI, NIDA, NIGMS, and NINDS) who provided guidance on application and submission processes. Each leader provided guidance and helpful application and submission information represented by the institutes in attendance and beyond. The webinar recording and resources page posted to our webpage as direct diversity supplement resources and within about a month following we released our repository first edition and a “submit request for access” button on the same webpage.

#### Match making service

Our webpage resource for diversity supplements offers a matchmaking service for investigators or trainees who would like to collaborate on proposals. When the “submit interest” button is selected a new email automatically generates addressing our team. Interested PIs or trainees then detail their inquiry, and our team assesses feasibility and provides guidance.

#### Raising Awareness

Our Program Manager, who communicates with our program scholars, their mentors and other academic and staff affiliates within and outside of our institution, includes a link to our Diversity Supplement Resources webpage as part of her email signature line. In particular, high volume communication periods with professors and mentor faculty such as during K program application review and grant cycle renewal periods spreads the word about our initiative and coincides with requestors finding out about our repository.

#### Funding opportunities list serv

ACTRI promotes repository access through monthly career development digests sent to KL2 scholars (both funded and ACTRI-supported) and Pilots Program awardees. These digests include comprehensive resources: funding opportunities, training events, workshops, and access instructions for both UCSD and UCLA grant repositories. Through the UC system and CTSA hub collaboration, UCSD scholars can access UCLA’s expanded grants library, which includes successful diversity supplements, R and K grants, without requiring UCLA affiliation.

#### Newsletter stories/Blog posts

A communications specialist at the ACTRI developed a blogpost series, which featured the diversity supplement initiative. We nominated one of the repository investigator contributors as an interviewee. The story highlighted both perspectives of the PI and the student on their processes of applying for a diversity supplement together and what fueled their mentor-mentee partnership to achieve that goal. Additionally, we promoted this initiative to announce the launch of our diversity supplement repository access, by including a featured segment in the quarterly ACTRI newsletter describing the resource and our new webpage to learn more.

### UCSD Hub-Linked Diversity Supplement Submission

To model engagement in Diversity Supplements, we have prioritized diversity supplement applications to the accompanying parent U award that supports ACTRI. With a fully enrolled KL2 program (8 slots) and over two years of remaining funding, we pursued diversity supplement opportunities through NCATS in 2021-2022, resulting in one funded application in 2023 out of three submissions. We support applicants by providing access to our supplement repository for guidance and offering comprehensive administrative assistance from both PI and finance teams. Additionally, we encourage KL2 program applicants to consider diversity supplements and institutional programs targeting underrepresented scientist retention.

## RESULTS

### Repository

The repository has grown since its launch in 2021 to over 51 diversity supplement submissions in 2024. Of 135 PIs that we contacted, 47 (35%) submitted successful diversity supplements. Seven PIs contributed more than one successful grant, comprising 16% of the sample. As seen in Table 1, PIs demographics showed equal number of men and women. Fourteen NIH Institutes are represented, with the four most prevalent being NIDA, NIGMS, NINDS, and NIA. More than half of PIs/awardees consented to sharing outside of UCSD.

**Table 1:** Diversity Supplement Repository.

| **Characteristic** | **Frequency and Percent (n=51)<sup>1</sup>** |
| --- | --- |
| **Year Awarded, supplements received (%)** |  |
| 2002-2016 | 6 (12%) |
| 2017-2019 | 13 (25%) |
| 2020-2022 | 20 (39%) |
| 2023-2024 | 12 (24%) |
| **PI Gender, n (%)** |  |
| Female | 21 (50%) |
| Male | 21 (50%) |
| **Agency, n (%)** |  |
| NIH | 50 (98%) |
| NSF | 1 (2%) |
| **NIH/NSF Institute, n (%)** |  |
| NIDA | 11 (22%) |
| NIGMS | 9 (18%) |
| NINDS | 7 (14%) |
| NIA | 6 (12%) |
| NIAAA | 4 (8%) |
| NIMH | 3 (6%) |
| NCI | 2 (4%) |
| NHLBI | 2 (4%) |
| NICHD | 2 (4%) |
| MCB (NSF) | 1 (2%) |
| NCATS | 1 (2%) |
| NIAID | 1 (2%) |
| NIAMS | 1 (2%) |
| OD | 1 (2%) |
| **Level Awardee, n (%)** |  |
| Undergraduate | 2 (4%) |
| Postbaccalaureate | 6 (12%) |
| Graduate | 20 (39%) |
| Postdoctoral | 23 (45%) |
| **Sharing preference, n (%)<sup>2</sup>** |  |
| UCSD only | 21 (44%) |
| Broader than UCSD only | 27 (56%) |

<sup>1</sup> Note that othere were 42 total PIs due to repeat sharing<sup>2</sup> UCLA provided 3 approved supplements for broad
sharing

#### Repository Access Analysis

Potential applicants of diversity supplement awards gained access to our UCSD repository from 2021-2024. Of the 51 repository users, the majority were affiliated with UCSD Health (53%), followed by UCSD Campus (24%). The remaining users included joint UCSD/SDSU affiliates (12%), dual UCSD Health/Campus affiliates (8%), and single users from SDSU and other affiliations (2% each). Numerous ways of hearing about the UCSD diversity supplement repository are evident by the selections users reported: UCSD affiliate/contact (N=16), ACTRI website (N=9), word of mouth (N=9), other or other website (N=5), newsletter/bulletin (N=2), NIH webinar (N=1) and ACTRI staff (N=1).

In evaluating time-based trends, we have observed upticks during seasonal periods of higher email volume as our team communications increase when requesting potential faculty reviewers for KL2 application periods and potential faculty mentors for the NCATS K12 program grant application. As shown in Figure 1, there was a steady rate of requests for access (35 requests for access in 15 months) from the announcement and launch in June 2021 until September 2022, followed by a lower rate of requests through 2023 (Figure 2).

**Figure 1.**
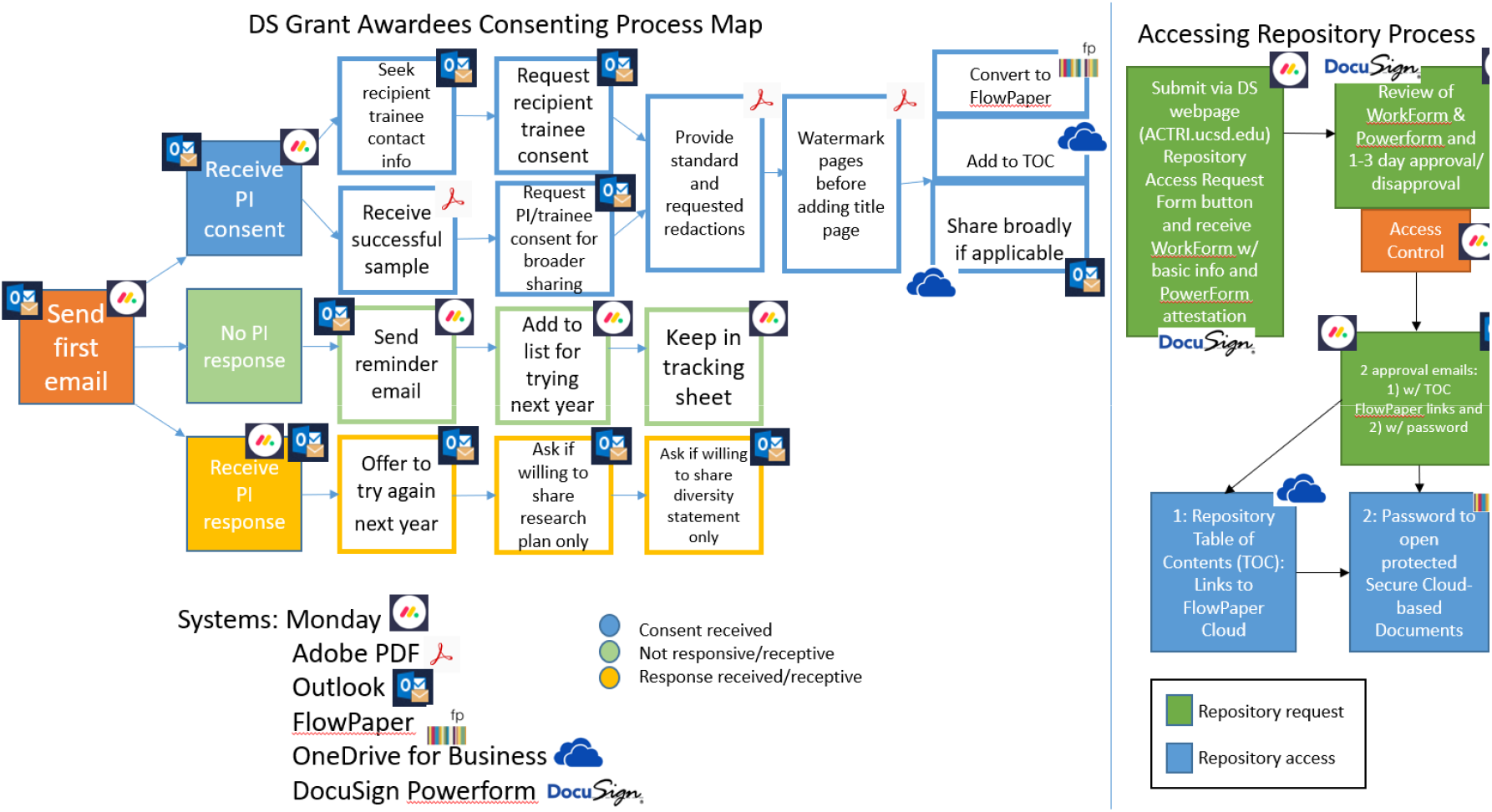
Consent outreach workflow to share Diversity Supplement PIs’ award information and Process Map for setting up repository access.

**Figure 2.**
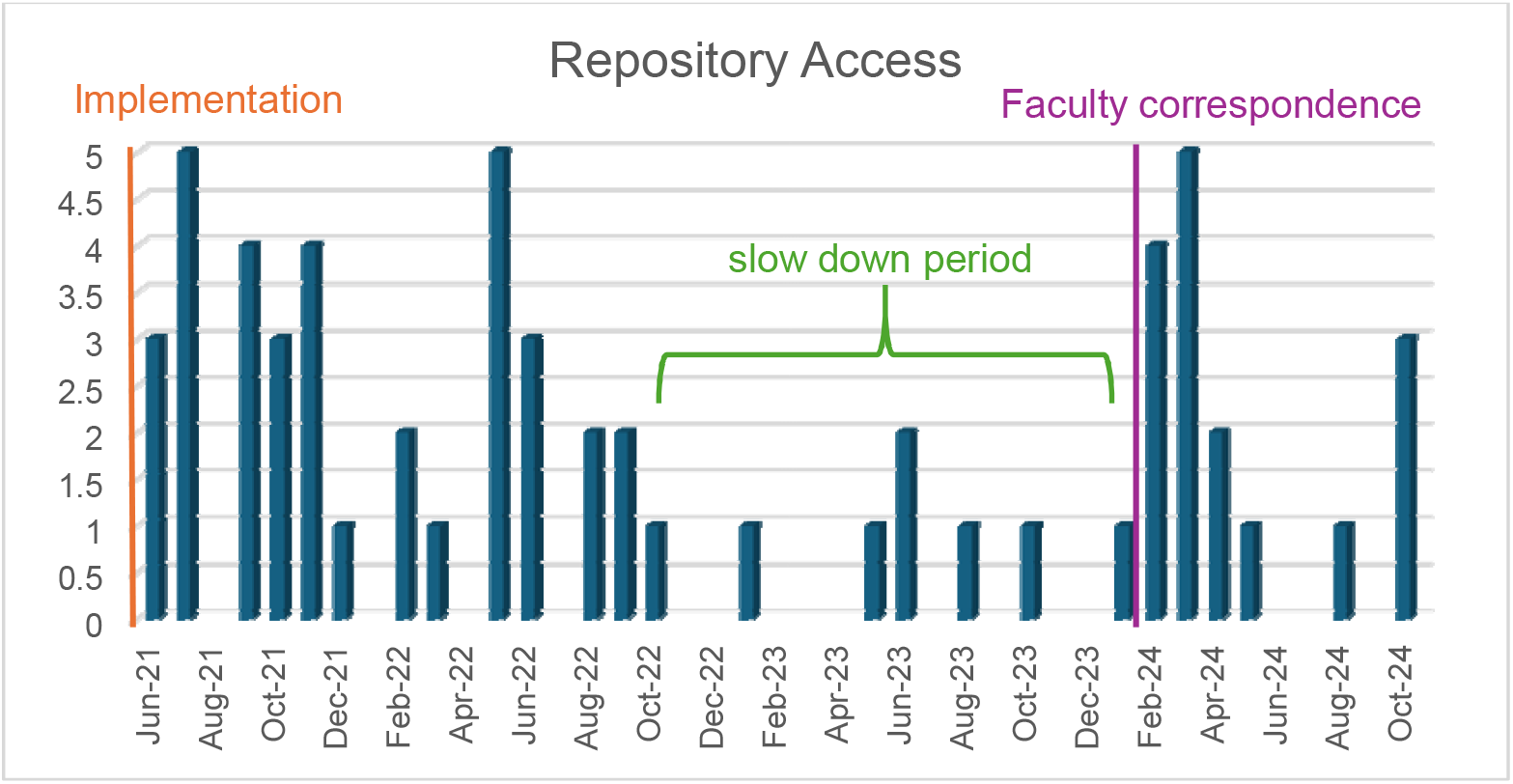
Trends of repository requests for access since implementation in 2021.

Towards the end of 2023 UCSD ACTRI had a call for KL2 reviewers during our application period followed by requests for K12 faculty mentors starting in January 2024 during which time data shows a significant increase in repository access requests steadily coming during those 5-months, January to May 2024, and averaging 2.6 per month.

#### Matchmaking

The matchmaking service, launched in June 2021 alongside the repository, has seen limited engagement. Since launch, we received only eight total inquiries: two from potential trainees (one student in November 2022 and one postbaccalaureate staff in May 2024) and six from eligible PIs. Most PIs sought postdoctoral trainees, but identifying eligible candidates proved challenging due to limited demographic and socioeconomic data from Human Resources and few postdoctoral trainees meeting diversity criteria.

### Trends in Supplements at UCSD

The number of diversity supplement grants awarded at UCSD pre-implementation (2013-2021) was 11 per year on average. Our post intervention average per year (2021-2023) is 21, up by 10.

## DISCUSSION

The underuse of NIH Diversity Supplements represents a significant missed opportunity to advance workforce diversity in biomedical research (3). In alignment with David et al. (1) recommendations for strengthening institutional administrative support, UCSD ACTRI established an infrastructure to support diversity supplement applications. To date, we have amassed a repository of over 50 successful grants that span 14 different NIH and NSF institutes, and this resource has been accessed by over 50 investigators and scholars. Moreover, ACTRI developed a comprehensive outreach strategy featuring informational webinars, success stories shared through social media channels, and clear pathways for connecting potential mentees with interested mentors. This systematic approach to diversity supplement implementation aligns with best practices identified in the literature while providing practical solutions for the specific needs of the UCSD research community. Since this resource was established, our institution doubled the rate of diversity supplements awarded. Although we do not know if this effort will continue to be as successful in the long-term, we are optimistic that the resources will continue to impact awareness, submissions, and receipt of these supplements and ultimately increase diversity the biomedical workforce.

### Lessons Learned

Recognizing that our experiences and resources may not generalize to other institutions, some of the lessons learned in this process may be useful to other hubs seeking to establish programs to stimulate diversity supplements. We outline these below:

1. **Tracking Supplements:** A key first step to improving the number of diversity supplements is finding and tracking awarded supplements. Doing so was challenging through public-facing databases (e.g., NIH Reporter), since not all supplements are diversity supplements and not all diversity supplements are listed under a specific FOA. Similar if not greater challenges were present for non-NIH supported supplements (e.g., NSF). Paralleling FAIR principles of data curation (8), making diversity supplements more findable would support future efforts.
2. **Learning from and Collaborating with Other Institutions:** We benefitted greatly from learning from best practices and processes established at other CTSA hubs, and we also established a process for cross-hub sharing of successful repository grants for PIs and recipients who consented to sharing. Encouragingly, more than 50% of PIs/recipients consented to sharing beyond our institution. Given the potency of the CTSA network, even more formalized coordination amongst programs could accelerate diversity supplements as well as establish more centralized outcome tracking and post-award networking/resources.
3. **Learning from Faculty Members:** A total of 13% of PIs engaged for the repository had received multiple diversity supplements. Learning from and celebrating such PIs through surveys of best practices and through featuring their scholars and mentorship stories (e.g., newsletters) could provide role models for other faculty. Such PIs seem to have made applications for such supplements a part of their laboratory and academic lives, could provide additional insights in processes to support diversity supplements, such recruitment and mentoring.
4. **Engaging with multiple NIH institutes:** The timing, format, eligibility and process for diversity supplements varies considerably across NIH Institutes. Connecting with multiple program officers and leaders at NIH and coalescing conjoint workshops enabled highlighting common and unique aspects of supplements specific to Institutes.
5. **Raising Awareness through Multiple Channels:** Review of data on how resource accessors found out about our resources indicated that no single outreach approach stood above others. We also observed periods of high communication with PIs (e.g., during application cycles for KL2 opportunities) incidentally raised awareness. Therefore, broad, frequent, and multi-channel outreach approaches may be required to sustain awareness and intentions to submit diversity supplements.
6. **Matchmaking:** Our success with supporting connections among potential PIs and diversity supplement recipients has been limited to date. Two barriers that we encountered have been that outreach to potential recipients may require quite different channels than that typically used to reach faculty (e.g., emails; listservs) and that PIs seeking “matches” were more interested in advanced scholars than the pool of potential recipients. Best practices for matchmaking deserve further study.

### Limitations and Future Directions

These lessons learned should be considered in tandem with limitations, as our description above reflects one institution’s experience and may not generalize. Additionally, the ultimate goal is to increase the number of supplements awarded, and we cannot directly attribute increases seen at UCSD in diversity supplements to our resources or elements. Nonetheless, we have been highly encouraged to date by both participation in both contributing to and accessing our resources. Our Hub is actively working on enhancements to our resources, including a) post-award support and networking for supplement awardees and outcome tracking alongside additional CTSA Hubs, b) workshops to support supplement submission for applicants, and c) expansion of the grant repository to be inclusive of additional federal and non-federal opportunities (e.g., Harold Amos Foundation, Veterans Affairs). It is our hope that our experiences may be useful for other institutions seeking to stimulate diversity supplements as part of broader effort to diversify the biomedical workforce.

## Data Availability

All data produced in the present study are available upon reasonable request to the authors

## ACKNOWLEDMENT

We would like to thank Medardo Gaytan, Kevin Muschter, Jennifer Beaudette for helping us along this project.

We would also like to thank Lisa Chan UCLA and Vince Caiozzo and Brooke Piercy at UCI for their support during the development of the repository.

The authors declare no competing interests that could have appeared to influence the work reported in this manuscript.

## AUTHOR CONTRIBUTIONS

Maryam Gholami contributed to interpretation of data and analyses, the design and draft of the manuscript. Eva Kintzer contributed to collection and analyses of data, analyses of tools, implementation of the project, and drafting the manuscript. Michelle Wong contributed to the design of the work and draft of the manuscript. Davey Smith contributed to the design of the work and draft of the manuscript. Colin Depp contributed to the design of the manuscript, design and implementation of the project, interpretation of the analyses, and drafting the manuscript.

